# Clinical application of whole-genome sequencing for precision oncology of solid tumors

**DOI:** 10.1101/2024.02.08.24302488

**Authors:** Ryul Kim, Seokhwi Kim, Brian Baek-Lok Oh, Woo Sik Yu, Chang Woo Kim, Hoon Hur, Sang-Yong Son, Min Jae Yang, Dae Sung Cho, Taeyang Ha, Subin Heo, Jeon Yeob Jang, Jae Sung Yun, Kyu-Sung Kwack, Jai Keun Kim, Jimi Huh, Sun Gyo Lim, Sang-Uk Han, Hyun Woo Lee, Ji Eun Park, Chul-Ho Kim, Jin Roh, Young Wha Koh, Dakeun Lee, Jang-Hee Kim, Gil Ho Lee, Choong-Kyun Noh, Yun Jung Jung, Ji Won Park, Seungsoo Sheen, Mi Sun Ahn, Yong Won Choi, Tae-Hwan Kim, Seok Yun Kang, Jin-Hyuk Choi, Soo Yeon Baek, Kee Myung Lee, Sun Il Kim, Sung Hyun Noh, Se-Hyuk Kim, Hyemin Hwang, Eunjung Joo, Shinjung Lee, Jong-Yeon Shin, Ji-Young Yun, Junggil Park, Kijong Yi, Youngoh Kwon, Won-Chul Lee, Hansol Park, Joonoh Lim, Boram Yi, Jaemo Koo, June-Young Koh, Sangmoon Lee, Yuna Lee, Bo-Rahm Lee, Erin Connolly-Strong, Young Seok Ju, Minsuk Kwon

## Abstract

Genomic alterations in tumors play a pivotal role in determining their clinical trajectory and responsiveness to treatment. While targeted panel sequencing (**TPS**) has been a key clinical tool over the past decade, advancements in sequencing costs and bioinformatics have now made whole-genome sequencing (**WGS**) a feasible single-assay approach for almost all cancer genomes in clinical settings. This paper reports on the findings of a prospective, single-center study exploring the real-world clinical utility of WGS (tumor and matched normal tissues) with two primary objectives: 1) assessing actionability for therapeutic options, and 2) providing clarity for clinical questions. Of the 120 various solid cancer patients enrolled, 95 (79%) successfully received genomics reports within a median of 11 working days from sampling to report. Analysis of these 95 WGS reports revealed that 72% (68/95) yielded clinically relevant insights, with 69% (55/79) pertaining to therapeutic actionability, and 81% (13/16) to clinical clarity. These benefits encompass selection of informed therapeutics and/or active clinical trials with driver mutations, tumor mutational burden (**TMB**) and mutational signatures, pathogenic germline variants that warrant genetic counseling, and information helpful for inferring cancer origin. Our findings highlight the potential of WGS as a comprehensive tool in precision oncology and advocate for its integration into routine clinical practice to provide a complete genomic landscape for tailored cancer management.

## Introduction

Personalized medicine aims to customize cancer treatment strategies for each individual ensuring interventions that yield the best patient outcomes. Central to this approach is molecular profiling of tumors. Molecular profiling has established itself as an essential tool in clinical practice, identifying targetable alterations ^1^. Projects such as The Cancer Genome Atlas (**TCGA**) and International Cancer Genome Consortium (**ICGC**) have unveiled both prevalent and rare genomic anomalies ^2,3^, with many presenting actionable clinical implications. These genetic deviations not only suggest viable treatment paths but also provide insights into the severity of the disease, assisting physicians in customizing treatment plans ^4^. For instance, the presence of *ERBB2* (HER2) amplification in breast cancers is associated with more aggressive tumor characteristics and a poorer prognosis ^5^. However, the introduction of HER2 inhibitors, such as trastuzumab, has markedly enhanced patient outcomes in these scenarios ^6^. This exemplifies the critical role of identifying specific molecular alterations, like HER2 status in breast cancer, as a cornerstone for informed clinical decision-making in oncology. To this end, the FDA recognizes many conventional molecular diagnostic techniques, including immunohistochemistry, in situ hybridization, and conventional DNA sequencing ^7^.

More recently, next generation sequencing (**NGS**) techniques have emerged as more efficient and comprehensive molecular diagnostics as they can detect various genetic alterations at once from single tests ^8^. Indeed, NGS-based comprehensive genomic profiling (**CGP**) is rapidly transitioning into a standard, go-to technique in clinical practice, highlighted by its incorporation into the European Society for Medical Oncology (**ESMO**) guidelines ^9^. In this context, the NGS technique is recommended as an alternative to conventional testing methods for detection of specific actionable mutations in advanced non-squamous non-small cell lung, prostate, ovarian, cholangiocarcinoma and colon cancer ^9^. Furthermore, the NGS is recommended for evaluating tumor mutational burden (**TMB**), a molecular marker for the response to pembrolizumab (immune checkpoint inhibitor) in a few cancer types (cervical, specific neuroendocrine, salivary, thyroid, and vulvar cancers) ^9^.

Until recently, targeted panel sequencing (**TPS**) platforms have been the predominant methods for the CGP in clinical settings. TPS conventionally focuses on particular exons and introns of 50-500 known cancer genes (typically representing 0.01%-0.1% of the genome), While TPS offers advantages such as minimal sequencing throughput (approximately 1-10 Gb) and simple bioinformatic processes, it faces several limitations. Notably, TPS exhibits reduced detection sensitivity for off-target driver mutations, complex genomic rearrangements and mutational signatures ^10,11^. This narrow coverage, paired with delay in update of the panel design, might overlook emerging data and new actionable markers not on the panel. Because TPS does not typically assess matched normal tissues, differentiation of somatically acquired mutations from germline polymorphisms is often challenging ^12^.

These considerations underscore potential utility of whole-genome sequencing (**WGS**) in the clinic. WGS provides comprehensive information into somatic point mutations, copy number alterations (**CNAs**), and structural variations (**SVs**), proving essential for precision oncology and individualized treatment plans ^13^. As technology improves and becomes more affordable, incorporating WGS in standard clinical care is vital to fully realize genomic medicine’s potential in oncology.

A fundamental query regarding WGS concerns its practical applicability in real-world clinical scenarios. Some believe that the sheer amount of data from WGS might compromise its effectiveness in clinical settings. This research assessed the practical use of a commercially accessible WGS bioinformatics pipeline for cancer patients at Ajou University Hospital in Suwon, South Korea.

## Materials and Methods

### Study Design and Participants

The research protocol was approved by the Institutional Review Board (**IRB**) of Ajou University Hospital (Suwon, Korea; IRB Code: AJOUIRB-SMP-2022-278) and was carried out in alignment with the principles of the Declaration of Helsinki and the stipulations of Good Clinical Practice Guidelines. This prospective, single-center-based genomics trial in the clinical setting (Clinical Research Information Service registration: #KCT0008118) included adult participants with diverse solid cancer types and stages, conducted at Ajou University Hospital (Suwon, South Korea) from September 2022 to April 2023. Prior to their inclusion, informed consent was obtained and documented for all participants. The primary objective of the study was to assess the clinical utility of implementation of WGS into a real-world hospital setting. Inclusion criteria consisted of adult patients (age ≥ 19) with histopathological verified solid cancers, who were eligible for biopsy or surgical excision as part of their routine care regimen. Patients were enrolled if medical oncologists wanted to apply whole-genome sequencing for providing 1) actionability for therapeutic options, and/or 2) clarity for clinical questions. For the trial, a total of 120 CancerVision^TM^ assays (Genome Insight, San Diego CA, USA) were provided from Genome Insight (San Diego CA, USA). Patients were excluded if they had insufficient tissue samples or declined genetic testing. For mentioning each case, we used anonymized case IDs which cannot identify patients.

### Sequencing and detection of genomic variances

Clinical-grade WGS was performed using the WGS component of CancerVision^TM^ assay as previously reported ^12^. Briefly, WGS was performed on tumor samples obtained as part of routine clinical care either via surgery or biopsy and stored as fresh frozen (**FF**) tissue. For biopsy sample cores were retrieved first for routine pathology, followed by at least one additional core for cancer WGS. If acquisition of sufficient fresh cancer tissue was not possible, formalin-fixed paraffin-embedded (**FFPE**) tissues for pathology-reviews were alternatively obtained to extract cancer DNA. For the matched normal samples peripheral blood was used. DNA extraction and library preparation was performed at the Genome Insight Inc., in a Clinical Laboratory Improvement Amendments (**CLIA**)-certified laboratory. For DNA extraction, we used the Allprep DNA/RNA Mini Kit (Qiagen, Hilden Germany) and Allprep DNA/RNA FFPE Kit (Qiagen, Hilden Germany) for fresh and FFPE samples, respectively. For library preparation, we used TruSeq DNA PCR-Free (Illumina, San Diego, CA), and TruSeq DNA Nano (Illumina, San Diego, CA) library preparation kits for fresh and FFPE samples, respectively. Sequencing was performed on the Illumina NovaSeq6000 platform (Illumina, San Diego, CA) with an average depth of coverage of 40x for tumor and 20x for blood.

### Genomic Analysis and Interpretation

Comprehensive genomic analysis and interpretation were conducted using the CancerVision™ platform (Genome Insight, San Diego, CA, USA). Briefly, sequencing data were mapped to the human reference genome GRCh38 through a machine-learning-augmented burrows-wheeler aligner maximal exact matches algorithm ^12,14^. This was followed by preprocessing, which included marking duplicates and creating analysis-ready binary alignment map files. Base substitution and short indel calls were initially made using Mutect2 ^15^ and Strelka2 ^16^. The intersecting variant calls from both tools were consolidated. Tumor characteristics such as tumor cell fraction (**TCF**), tumor cell ploidy, and segmented CNA profiles were estimated via the Sequenza^17^, while Delly was employed for identifying somatic genomic rearrangements ^18^. For curating a high-confidence somatic variant dataset, we excluded variants with specific attributes: low mapping quality (Q<40), excessive indels or clipping (>90%), variant reads with more than five mismatched bases, and allele frequency of 1% or higher in a panel of normal samples. Rearrangements with suboptimal mapping quality (Q<25), inadequate supporting reads (n<5), or high discordance in matched normal samples were filtered out. Additionally, suspected library artifacts, like small deletions/duplications (<1Kbp) lacking soft-clipped reads, and unbalanced inversions (<5Kbp) without adequate supporting reads (n<5) were also excluded. The refined somatic variants underwent annotation via the Ensembl Variant Effect Predictor ^19^. Their therapeutic relevance was ascertained through external databases from the Catalog Of Somatic Mutations In Cancer (**COSMIC**) ^20^ and OncoKB ^21^. Germline polymorphisms were also annotated with the Ensemble Variant Effect Predictor to determine their pathogenicity^19^.

### Correction of FFPE artifacts

To remove sequencing artifacts abundant in FFPE tumor specimens ^22^, we instituted specialized point-mutation and CNA filters, harnessing machine learning (**ML**) methodologies. Briefly, we fashioned models based on features derived from the WGS data. Each was trained on FFPE samples, equipped with designated variant labels. In the realm of CNA assessments, we utilized a bespoke Genome Insight patented FFPE CNV rectification algorithm.

### Mutational Signature Analysis

Using the COSMIC database’s cataloged signatures, we assessed the prevalence of mutational signatures for single-base substitutions (**SBSs**), indels, and structural variations (**SVs**) in every sample, employing the non-negative least squares approach based on algorithms previously reported ^23^. The selection of signature sets was tailored to each sample based on the cancer type ^23^. For instances where the default signature combination did not sufficiently explain samples (cosine similarity < 90%), a manual evaluation was undertaken to adjust the signature set by either addition or removal.

### Analysis of Homologous Recombination Deficiency (HRD) and Microsatellite Instability (MSI)

To assess HRD, we developed our proprietary algorithm by combining HRD-associated features, such as mutational signatures of point mutations, and copy number changes ^24^. These included single-base substitution signatures (SBS3 and SBS8; reference signatures are available at https://cancer.sanger.ac.uk/signatures/), an indel signature (ID6), genomic rearrangement signatures (RS3 and RS5), deletions accompanied by microhomology, and CNVs. Custom scripts scaled scores of the multi-dimensional features using coefficients derived from published algorithms to compute the final HRD probability scores. Scores equal to or exceeding 0.7 were deemed HRD-positive. For a quantitative evaluation of somatic microsatellite alterations, we considered both the score from MSIsensor ^25^ and the proportion of MSI-related mutational signatures (SBS6, SBS15, SBS20, SBS21, SBS26, SBS30, SBS44) ^26^.

## Results

### Patients and characteristics

Between September 2022 and April 2023, the study initially enrolled 128 solid tumor patients, requested by medical oncologists in need of primarily two main questions: 1) assessing actionability for therapeutic options (referred to as actionability question; **Category I**), and 2) providing clarity for clinical questions (referred to as clarity question; **Category II**). In the study, 8 patients were excluded: 4 due to the absence of tumor specimens and 4 due to voluntary withdrawal (**Figure 1A**). The full demographics of the 120 patients finally enrolled in this study is shown in **Supplementary Table 1 (n=120)**.

**Figure 1.**
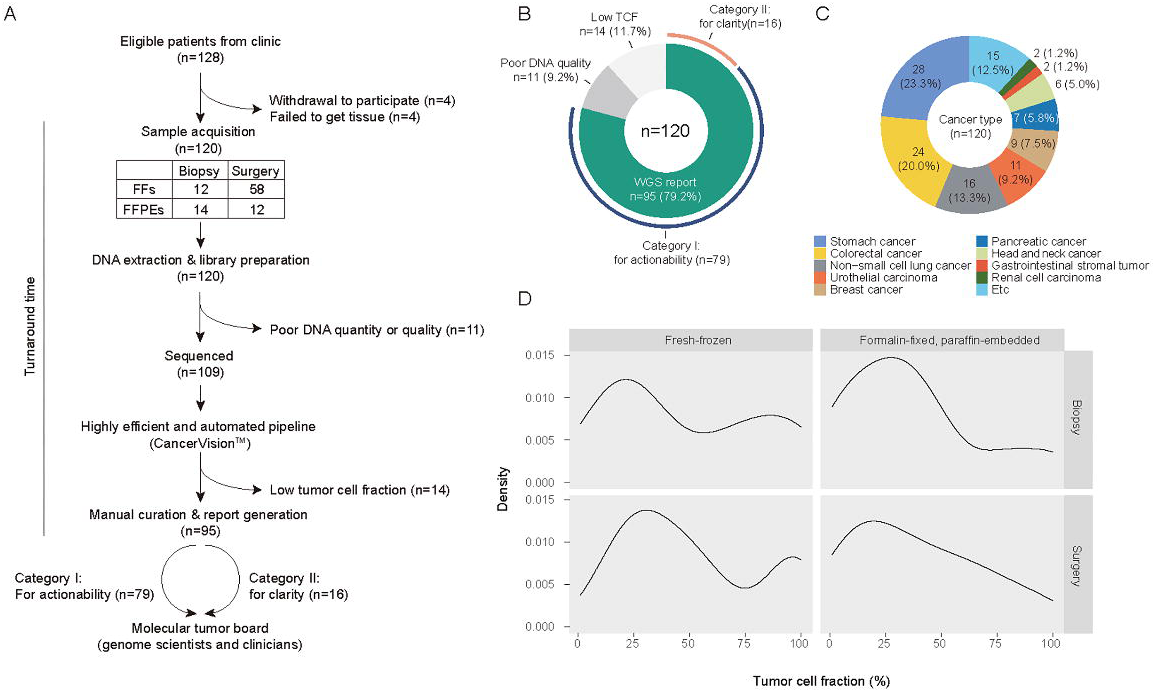
Study design and metrics of the sequencing cohort. **(A)** Flow diagram of the study protocol. **(B)** Technical success rate of CancerVision^TM^ application in the clinical setting. **(C)** Tumor types enrolled in this study. **(D)** The distribution of tumor cell fraction of cancer tissues estimated by WGS according to the specimen types.

Among the 120 patients finally enrolled, 11 of these were not sequenced due to suboptimal DNA quality/quantity extracted. Of the 109 patient’s genome sequenced, 14 yielded non-interpretable genome sequences attributed to a low tumor cell fraction (TCF <15%; **Figure 1A**). Consequently, interpretable whole-genome sequencing (WGS) reports were obtained for 95 patients, reflecting a 79% technical success rate (95/120) in the real-world application of WGS (**Figure 1B**)

The objective of this study is to observe the clinical application of WGS in a real-world setting, leading to a heterogeneous composition of our study cohort. Briefly, the entire cohort (n=120) included 23 tumor types (**Figure 1C**), consisting of 71 male (59.2%) and 49 female patients (40.8%) with a median age of 60 (range 19-85). At the point of sample acquisition, 99 (82.5%) were treatment-naïve, 7 (5.8%) were relapsed, and 14 (11.7%) were on active treatment. Tumor specimens were obtained from fresh-frozen surgery (n=58; 48.3%), fresh-frozen biopsy (n=36; 30.0%), FFPE surgery (n=12; 10.0%), and FFPE biopsy (n=14; 11.7%) (**Figure 1A**).

For the 109 patient’s genome sequenced, the median tumor cell fraction estimated from the genome sequences were on average 43.6% (range 0.0-100.0%). Contrary to the conventional wisdom, TCF was not substantially different between surgical and biopsy specimens (P=0.359 from Analysis of Variance; **Figure 1D**), suggesting widespread applicability of WGS regardless of the sample collection methods.

### Turn-around time in the clinical setting

For the 95 samples processed, the median turn-around-time (TAT), from sample acquisition to reporting, was 11 working days (range 9 to 18 days) (**Figure 2**). The breakdown of the duration included: lab experiments (median 2 days, range 2-15 days), pre-sequencing quality control and library pooling (median 5 days. range 1-8 days), sequence production (2 days), post-sequencing time (median 1 day, range 1-4 days), bioinformatics analysis (dry pipeline from raw fastq files to filtered mutation calls) a median 12.6 hrs (range 4.7-98 hrs); and report generation including final curation from medical genome scientists (∼1.5 hrs). In four cases, the library preparation step was substantially delayed due to unexpected breakdown of lab equipment. Overall, 87.4% of the patients (n=83) received their WGS reports in a two-week period. Considering 11-18 days of TATs for TPS in real-world settings ^27,28^, the TATs for WGS in our study were found to be generally satisfactory.

**Figure 2.**
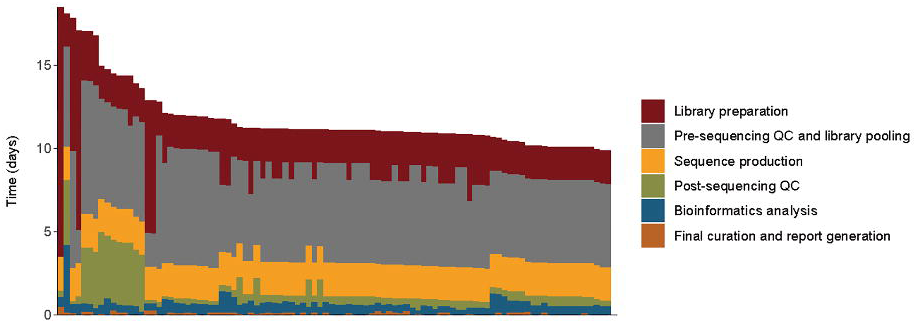
Turn-around time of the WGS-based cancer genome profiling in the clinical setting. TAT was measured in working days from sample collection to the acquisition of the WGS report. Processes of the WGS assay were subdivided into six components, including 1) library preparation, 2) pre-sequencing quality assessment and library pooling, 3) sequence production, 4) post-sequencing process, 5) bioinformatics analysis, and 6) final curation/report generation.

### Mutational detection

Our WGS analysis of 95 tumor/normal matched patient samples revealed a total of 3.54 million somatic mutations including 1,436,165 single-base substitutions (**SBSs**), 2,089,735 indels and 11,614 structural variations (**SVs**). The mutational burdens of SBSs were close to previously known burdens (**Figure 3A**), suggesting that our sequencing analyses were overall conducted properly. Mutations altering known cancer genes, such as *EGFR* L858R, *KRAS* G12V, and *BRAF* V600E, along with their mutational signatures and genome-wide CNAs were summarized in **Figure 3B**. Collectively, 288 CNA and 261 SV events altered known cancer genes (**Figure 3C**). For each sample, a combined mutational landscape was obtained (**Figure 3D)**, which cross validated to each other to evaluate the accuracy of the calls. Of note, these combined information are typically challenging to access in other assays. In our cancer cohort, 32 (33.7%) and 75 (62.5%) of the tumors carried whole-genome duplications (ploidy > 3.5) and complex rearrangements (including chromothripsis, chromoplexy and breakage-fusion-bridge cycles), respectively, which were associated with poorer drug responses and survivals ^29,30^. Beyond these somatic mutations, we identified 110 rare (<0.5% in population) pathogenic or likely-pathogenic germline variants in cancer predisposition genes (**Supplementary Table 2**).

**Figure 3.**
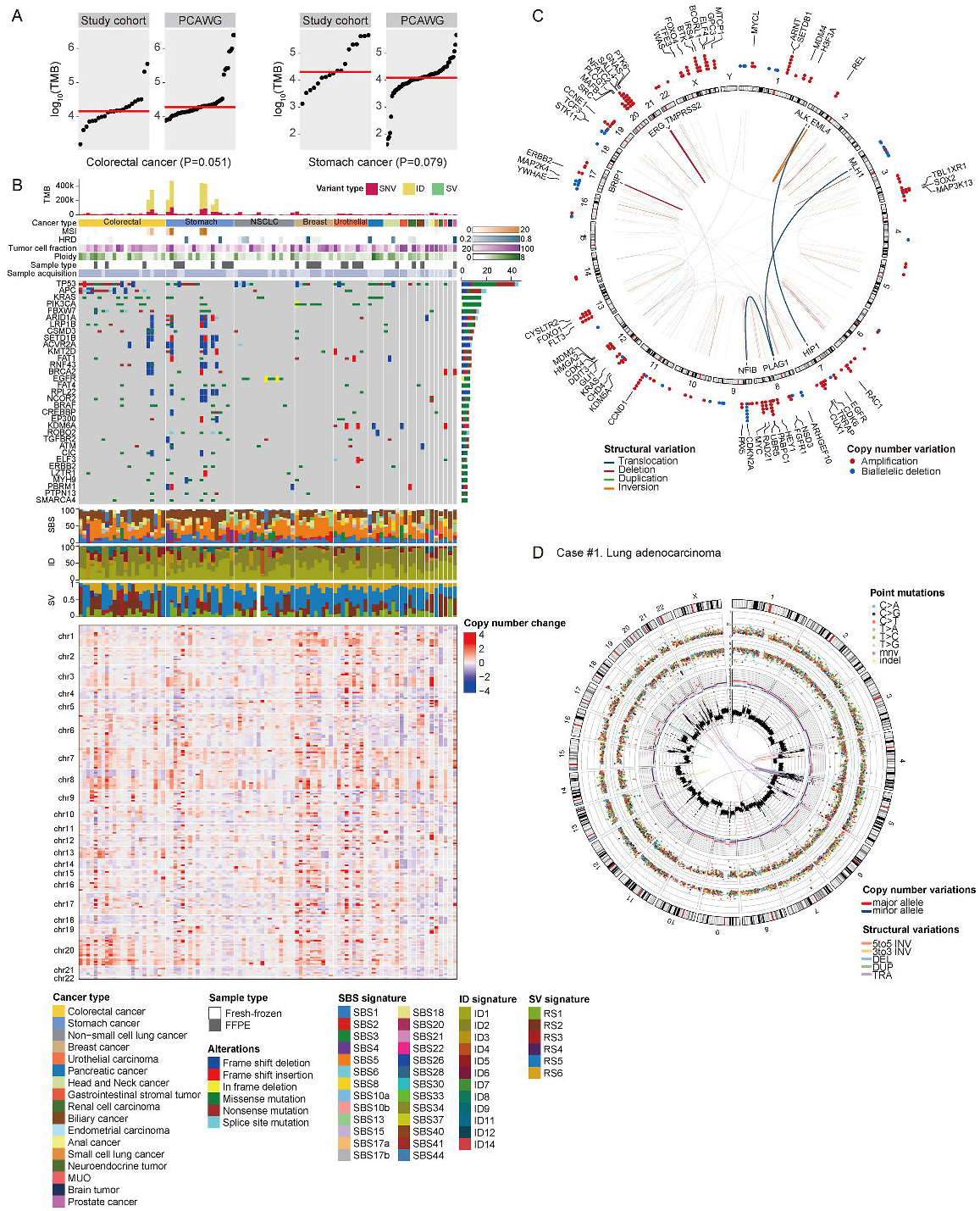
Landscapes of somatically acquired mutations in cancer genomes. **(A)** Comparison of tumor mutational burden (TMB) between PCAWG (Pan-Cancer Analysis of Whole-Genomes) and this study. Two studies showed similar TMBs in two cancer types. Every dot represents a sample, and the red horizontal lines are the median TMBs in the respective cancer type. **(B)** Landscape of somatically acquired mutations in 95 solid tumors in this study. Top to bottom: tumor mutational burden (TMB), cancer type, microsatellite-instability (MSI) score, homologous recombination deficiency (HRD) score, tumor cell fraction estimated from WGS, genome ploidy, sample type (fresh-frozen, or FFPE), sample acquisition method (biopsy, or surgery), point mutations in frequently mutated cancer genes, mutational signatures (SBS, ID, and SV), and copy number alterations. **(C)** Cancer genes frequently altered by SVs (arcs inside the circle) and copy number changes (dots outside the circle). (D) A representative circos plot which summarizes all the somatic mutations (from outer to inner circles; chromosomes, point mutations with variant allele fraction (VAF), point mutations with inter-mutational distance, genome-wide loss-of-heterozygosity (LOH) pattern, genome-wide copy number changes, and SVs)

### Actionability with approved therapeutics (I-1)

Of the 95 patients in the cohort, 79 were enrolled primarily for assessing therapeutic actionability. Among these, for 55 patients (70%), the findings from WGS were supportive to the clinical decision-making process (**Figures 3B, 4A**).

The actionable results from WGS were divided into three distinct subgroups: (1) informing the selection of targeted therapeutics (**Category I-1**; n=28; 35.4%), (2) facilitating the screening for mutation-specific clinical trials (**Category I-2**; n=22; 27.8%), and (3) aiding in the elimination of ineffective treatment options (**Category I-3**; n=5; 6.3%)(**Figure 4A**). The actionability was variable according to the tumor types, and the non-small cell lung cancer showed a higher chance of Category I-1 actionability (**Figure 4B**; P=0.360 by exact binomial test) although it did not reach statistical significance, likely due to the low number of samples enrolled. On the contrary, colorectal cancer showed lower Category I-1 chance than average (**Figure 4B**; P=0.630 by exact binomial test).

**Figure 4.**
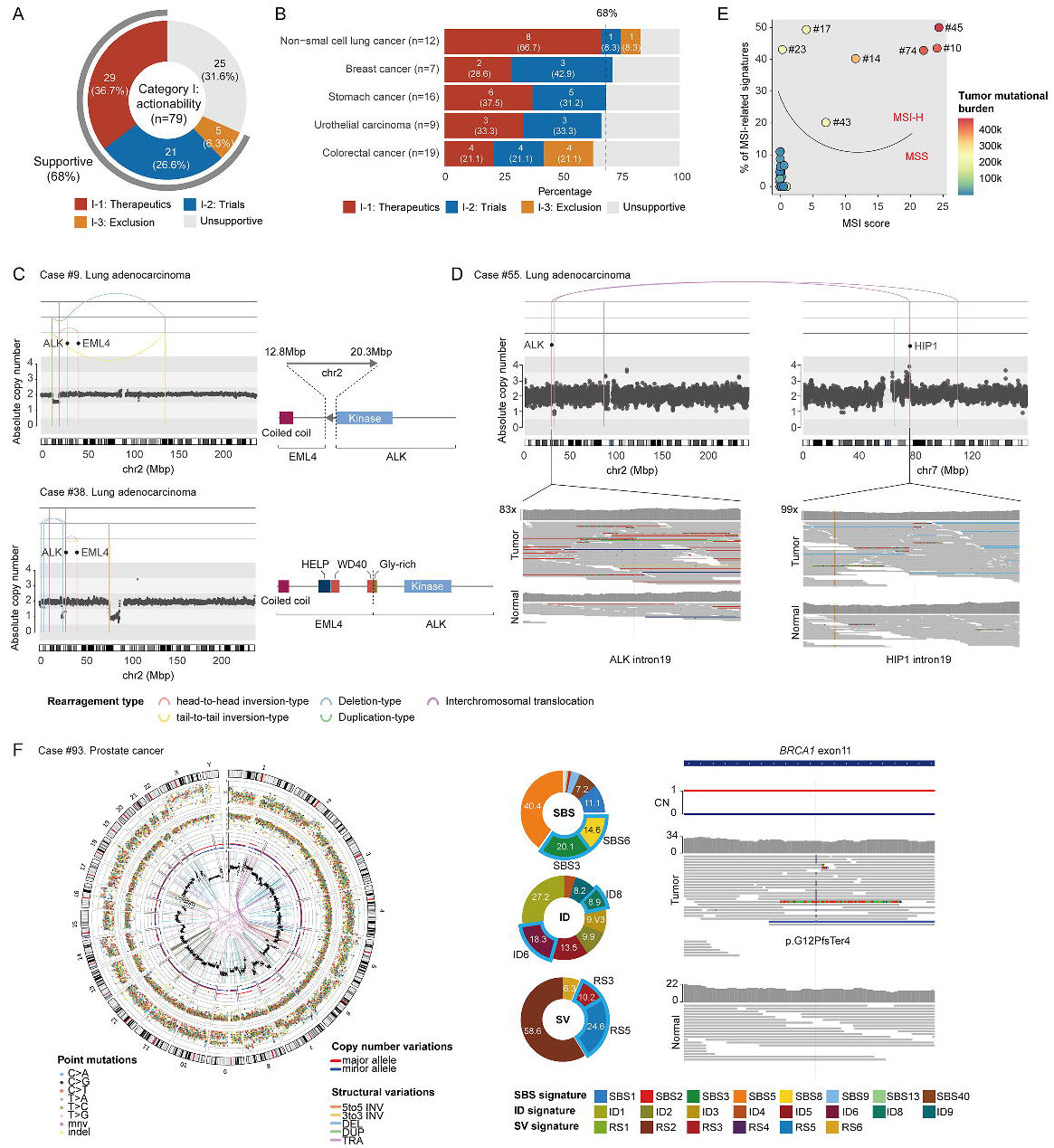
Clinical utility of WGS for assessing therapeutic options. **(A)** Overall, WGS was supportive in ∼70% of the cases conducted for therapeutic options. WGS provided information for therapeutics (I-1), clinical trials (I-2), or exclusion of options (I-3). **(B)** Clinical utility for assessing therapeutic options across cancer types. **(C)** Two lung adenocarcinoma cases harboring an oncogenic *EML4*-*ALK* fusion gene by balanced chromothripsis mechanisms. **(D)** A lung adenocarcinoma case harboring a rare *HIP1*-*ALK* fusion gene. **(E)** Seven cases of GI tract cancers showing the MSI-H feature, a mutational pattern-based target for pembrolizumab treatment. Two independent information, i.e., the MSI score and MSI-associated mutational signatures, clearly distinguishes MSI-H cancer samples. **(F)** A prostate cancer case with the HRD feature, a mutational pattern-based target for PARP inhibitors. A circos plot showing HRD feature, proportions of HRD-associated signatures for SBSs, indels, and SVs, and mutations induced HRD in the cancer (somatically acquired BRCA1 frameshift insertion and loss-of-heterozygosity events) are shown.

In **Category I-1** (n=28; **Supplementary Table 1**), 15 cases harbored actionable point mutations, such as *EGFR* L858R in lung adenocarcinomas (actionable with erlotinib and others) ^31^, *BRAF* V600E in colon cancers (actionable with encorafenib) ^32^, *PIK3CA* H1047R/N345K in hormone positive breast cancers (actionable with alpelisib) ^33^, and *FGFR3* G370C/S249C/Y737C in urothelial cancers (actionable with erdafitinib) ^34^. Often, relatively infrequent, but actionable point drivers were also captured, such as *PIK3CA* N345K in hormone positive breast cancer (actionable with alpelisib).

Frequently, we identified actionable targets beyond the simple point mutations. Three lung adenocarcinomas carried *ALK* fusion genes by SVs (all actionable with alectinib, brigatinib or lorlatinib) ^35–37^. In two of these (Cases #9, #38), balanced chromothripsis, a type of complex rearrangements, formed *EML4*-*ALK* fusion gene (**Figure 4C**) ^38^. In the remaining one (Case #55), translocations between chromosomes 2 and 7 shaped an atypical ALK fusion gene (*HIP1*-*ALK*; **Figure 4D**) ^39^. Additionally, seven gastrointestinal cancer cases (six stomach and one colon cancers) exhibited microsatellite instability high (**MSI-H**) features, indicating pembrolizumab as a potential primary therapeutic target ^40^. WGS offered dual insights into MSI-H (both the mutation burden at microsatellite loci and MSI-related mutational signatures), allowing for a clear differentiation of MSI-H phenotype cancers from MSS tumors (**Figure 4E**).

In **Category I-1**, two cases displayed actionable features more sensitively detected by WGS than by conventional methods. First, a prostate cancer case (Case #93) presented a clear homologous recombination deficiency (**HRD**) phenotype (**Figure 4F**; actionable with olaparib, niraparib, or talazoparib) ^41–43^. This was characterized from the combination of diverse somatic mutation profiles, including a substantial mutational burden of significant single-base substitutions (SBS3; n=1,452; 20.1% of all 7,232 SBSs), indels (ID6; n=142; 18.3% of all 827 indels), and genomic rearrangements (RS3 and RS5; n=107; 34.8% of all 323 SVs). Despite the HRD phenotype, WGS analysis determined the sporadic nature of the cancer, with two somatically acquired mutations: a frameshift insertion and a combined loss of heterozygosity in the *BRCA*2 gene (**Figure 4F**).

### Actionability with clinical trials (I-2)

As briefly mentioned above, a substantial portion of the cases had the potential for enrollment in clinical trials based on WGS findings (n=22 out of 79; 27.8%; **Category I-2**; **Figures 3B, 4A**; Information of possible trials are shown in **Supplementary Table 1**). Among these, 12 cases showed point mutations in key target genes, such as *KRAS* (n=4), *PIK3CA* (n=3), *HRAS* (n=2), and others (one for each of *BRAF*, *PTEN*, *ERBB2* and *AKT*). Additionally, six cases were potential candidates for clinical trials through their focal amplifications encompassing *CCND1* (n=3) and *MET* (n=1), or a fusion gene involving *NRG1* (**Figure 5A**).

**Figure 5.**
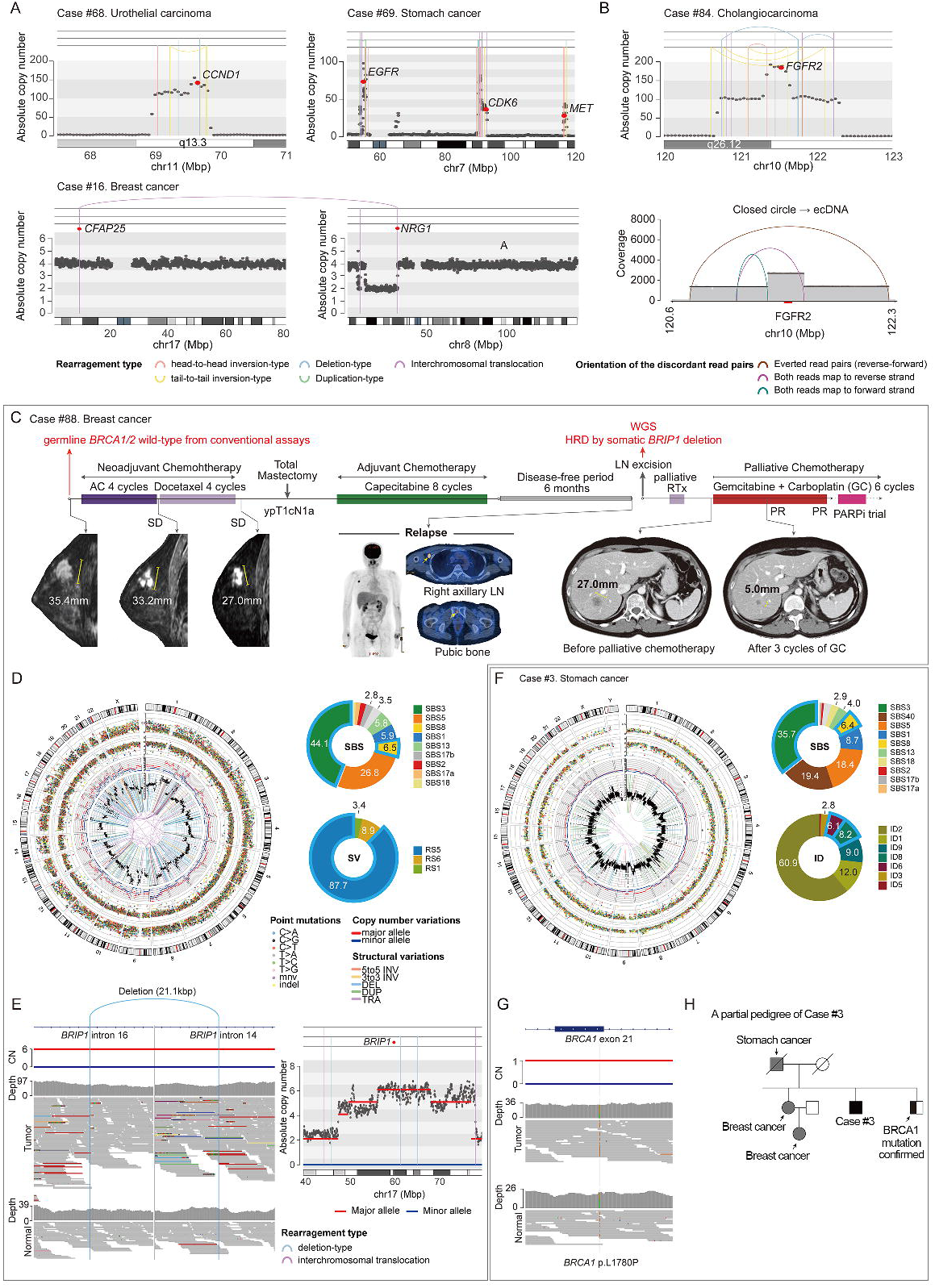
Clinical utility of WGS for assessing therapeutic options: available clinical trials (Category I-2) **(A)** Three cancer cases showing oncogene amplification (*CCND1*, *EGFR*) or *NRG1* rearrangement, which are genomic targets for clinical trials. **(B)** A cholangiocarcinoma case harboring ecDNA-mediated *FGFR2* hyper-amplification, a genomic target for a clinical trial. **(C)** The clinical course of a triple negative breast cancer patient who found a possible clinical trial from WGS. Surgically removed tissue from the right axillary lymph node (LN) underwent WGS. WGS identified the strong HRD feature and its underlying genetic cause (somatically acquired *BRIP1* complete inactivation), which are targets for a clinical trial. **(D)** Circos plot demonstrating characteristic HRD features of the patient. Outer to inner: ideogram, point mutations (single base substitutions [SBSs] and short indels) and their variant allele frequencies, distances between adjacent point mutations, major (red line) and minor (blue line) allelic copy number (CN), total segmented CN (black dot) and structural variations (SVs). SBS and SV mutational signatures associated with HRD are shown in pie graphs **(E)** Integrative genome viewer (IGV) snapshot of a somatic SV disrupting BRCA1 interacting helicase 1 (*BRIP1*) gene: 21.1Kbp deletion between *BRIP1* intron 14 and *BRIP1* intron 6. **(F)** A stomach cancer case who found a possible clinical trial from WGS. WGS identified the strong HRD feature and its underlying genetic cause (a germline BRCA1 pathogenic variant combined with loss-of-heterozygosity of the locus in the cancer), which are targets for a clinical trial. A circos plot showing the HRD feature as mentioned above, and two pie graphs show HRD-associated mutational signatures. **(H)** Pathogenic mutations underlying the HRD feature in the case: we identified germline *BRCA1* mutation with loss of heterozygosity in the tumor sample. **(H)** Pedigree of the patients. WGS enabled the patient to recognize a strong cancer predisposition in his family. The pathogenic BRCA1 p.L1780P variant was also identified in the younger sibling’s genome.

A cholangiocarcinoma patient (Case #84) exhibited focal, extra-chromosomal DNA (**ecDNA**)-driven *FGFR2* hyper-amplification ^44^, reaching approximately 185 copies (**Figure 5B**). While FGFR2 fusions or rearrangements are indicated a potential treatment with the FGFR-targeting tyrosine kinase inhibitor, pemigatinib, FGFR2 amplifications such as this patients are known to be less responsive to pemigatinib ^45^. An ongoing clinical trial (NCT04526106) is evaluating the efficacy of RLY-4008, a highly selective FGFR2 inhibitor, in patients with solid tumors with FGFR2 amplification.

In our cohort, HRD phenotype findings from WGS correlated with potential clinical trial opportunities for three patients. One case was particularly illustrative the insight gained from WGS of a female in her 40s with triple-negative breast cancer (Case #88; **Figure 5C**). Her young age at diagnosis initiated conventional *BRCA1/2* testing, which yielded a negative result. Given the information, she underwent neoadjuvant chemotherapy (adriamycin, cyclophosphamide, and docetaxel) with a limited response (stable disease from the Response Evaluation Criteria in Solid Tumors (RECIST v1.1) criteria). She experienced local recurrence in the right axillary lymph nodes and distant metastases in the pubic bone 6 months post mastectomy and adjuvant chemotherapy (8 cycles of capecitabine). Unexpectedly, WGS analysis of a specimen from the axillary lymph nodes revealed a strong HRD signature (HRD score = 0.924; **Figure 5D**). Consistent with the previous *BRCA1/2* testing, pathogenic germline mutations in *BRCA1* and *BRCA2* genes were not detected in WGS. However, the *BRIP1* (BRCA1-interacting helicase 1) gene was completely inactivated by two somatically acquired mutations (21.1 kbp-sized deletion between intron 14 and intron 16 and a combined loss of heterozygosity of *BRIP1* gene; Figure **5E**). This finding likely contributed to the observed HRD phenotype in her cancer ^46^. Informed by the WGS insights, the patient was prescribed a platinum-based treatment regimen, recognizing the connection between homologous recombination pathway deficiencies and therapeutic efficacy ^47^. After receiving gemcitabine plus carboplatin for three cycles, she exhibited a partial response, which continued through six cycles (**Figure 5C**). Presently, her suitability for a talazoparib (PARPi) maintenance therapy clinical trial (NCT04755868) is being considered, a decision steered by the WGS findings.

The remaining two cases (Cases #3, #76) were stomach cancers, HRD phenotypes of which were recognized by the strong HRD signatures in their somatic mutation profiles (0.718 and 0.840, respectively; Case #3 is represented in **Figure 5F**). These strong signatures suggested a familial origin for these cancers. Subsequent analysis for germline variants revealed causative mutations: a germline pathogenic variant in *BRCA1* p.L1780P for Case #3, and in *BRCA2* p.C717* for Case #76. Additionally, the secondary hits at these loci were observed in their matched cancer tissues (a large deletion for Case #3; a somatically acquired stop-gain base substitution for Case #76), reassuring the causative roles of the inherited mutations for development of the tumors (**Figure 5G**). These findings indicated the potential for enrollment in a PARP inhibitor clinical trial for any solid tumors with deleterious mutations in HRD-associated genes (NCT04171700). Importantly, the WGS results were the first to recognize their familial cancer history. Of note, WGS initiated the patients to first recognize their familial cancer history (**Figure 5H**), and to prompt genetic counseling for their family members. We further found the pathogenic germline mutation in his younger sibling of Case #3.

### Actionability by exclusion of non-beneficial therapeutic strategies (I-3)

In five patients, WGS assisted clinical decision making through informing exclusion of potentially ineffective therapeutic options (**Category I-3**). These include four colorectal cancers (two with *KRAS* G12V, one with *KRAS* Q61L, one with *NRAS* G61K; Cases #18, #28, #79 and #21; **Supplementary Table 1**), and one non-small cell lung cancer with no targetable mutations (Case #29). Although RAS oncogenic mutations were the earliest drivers discovered in human cancers, they have not been druggable for ∼40 years ^48^. Recently, a few *KRAS* G12C-selected inhibitors, such as sotorasib and adagrasib, were approved for the treatment of solid tumors ^49,50^. However, *KRAS* mutations other than G12C are still intractable for other targeted agents. These genomic mutations led clinicians to avoid potential targeted therapies, such as cetuximab in colorectal cancers ^51^ and erlotinib/crizotinib in lung adenocarcinoma ^52^.

### Clarity for clinical questions (II)

Of the 95 patients in the cohort, 16 were enrolled, not primarily for understanding therapeutic actionability, but for getting insights into the questions that are associated with clinical decision making (**Figure 1A**; **Supplementary Table 1**). These 16 patients were further categorized into three groups: (1) identification of drug resistance and/or responsive mechanisms (n=6; **Category II-1**), (2) resolution of tumor origin (n=7; **Category II-2**), and (3) evaluation of uncertain familial cancer cases (n=3; **Category II-3**). WGS successfully addressed these clinical questions in 81% (13/16) of the cases in the clarity category (**Figure 6A**).

**Figure 6.**
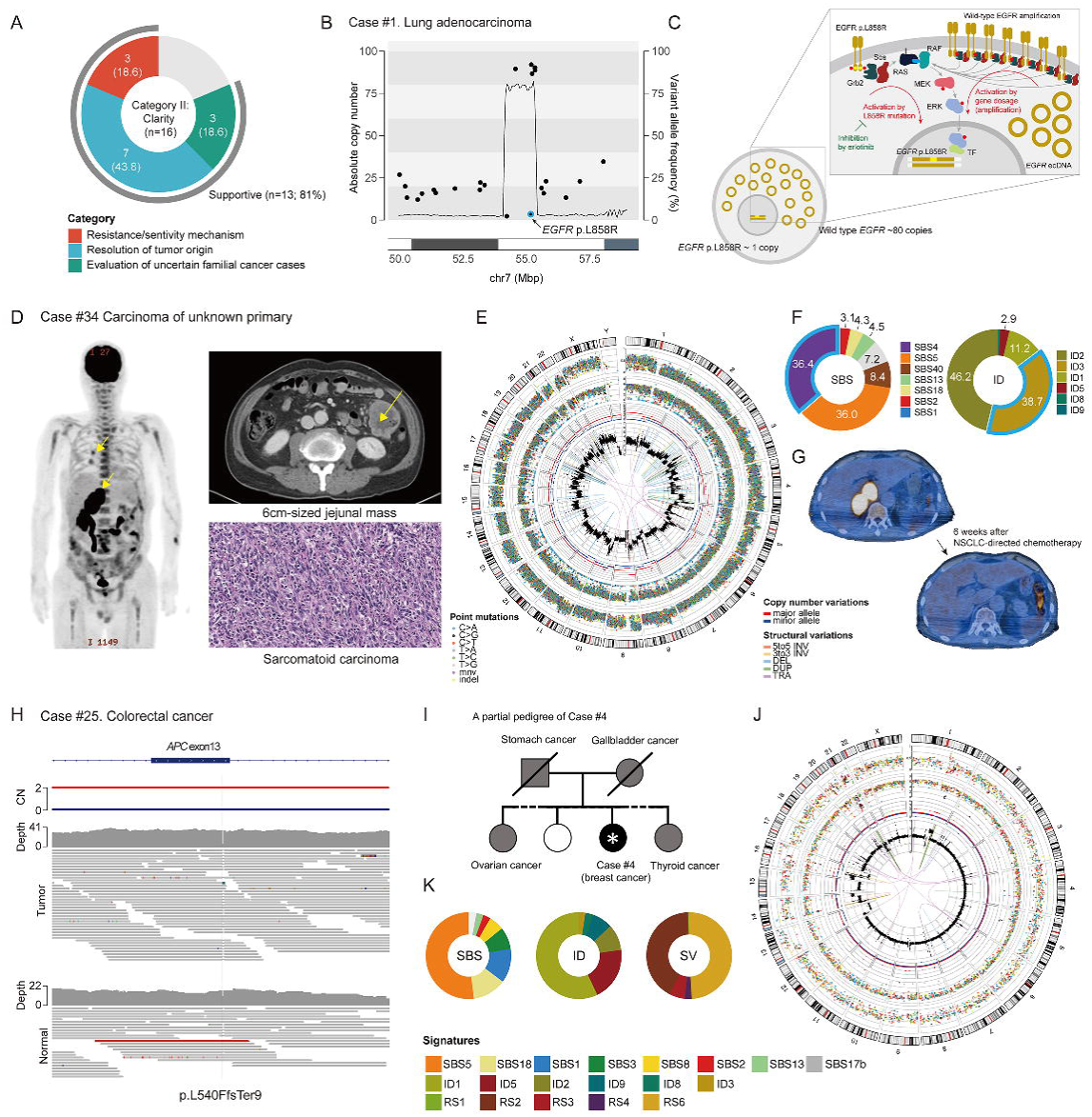
Clinical utility of WGS for providing clarity to clinical questions (Category II) **(A)** WGS was supportive in ∼81% of the cases conducted for clinical clarity. WGS provided information for resistance/sensitivity mechanism (II-1), tumor origin (II-2), or evaluation of familial cancer cases (II-3). **(B)** A lung adenocarcinoma case evaluated by WGS for understanding the resistance mechanism against erlotinib treatment. Hyper-amplification of the *EGFR* wild-type allele was found. **(C)** In the cancer, the MAPK pathway was activated by dual mechanisms: 1) EGFR L858R and 2) EGFR-wt amplification. The latter cannot be inhibited by erlotinib. **(D)** 2-deoxy-2-[fluorine-18]fluoro-D-glucose (18F-FDG) Positron-emission tomography-computed tomography (PET-CT) image at initial presentation of a patient with cancer of unknown primary site patient showing 6.4cm-sized jejunal mass. There was a hypermetabolic mass in the right adrenal gland with probable peritoneal carcinomatosis. Additionally, we noted multiple small nodules in both lungs. Computed tomography scan and hematoxylin and eosin image of the right adrenal gland mass are also shown. **(E)** Circos plot showing genomic findings from WGS. **(F)** Mutational signature analysis of SBSs and IDs. High proportions of SBS4 and ID3 (attributed to tobacco smoking) were noted. **(G)** Treatment response to non-small cell lung cancer (NSCLC)-directed chemotherapy: nivolumab plus ipilimumab in combination with paclitaxel plus carboplatin. **(H)** A colorectal cancer case with germline APC truncating mutation, suggesting the cancer was a familial case (familial adenomatous polyposis). **(I)** A bilateral breast cancer case suspected to be familial cancer given the clinical feature and familial history. **(J, K)** In the cancer genome profiling, HRD-associated features were absent, suggesting the bilateral breast cancer was likely sporadic. In line with the observation, no pathogenic germline mutations were found.

The six cases in the **II-1** subcategory include patients who were resistant (n=5) or hyper-responsive (n=1) to targeted therapeutics (**Supplementary Table 1**). Post-treatment cancer tissues were analyzed through WGS to understand the molecular basis of these unusual treatment sensitivities, yielding conclusive insights in half of the cases (3/6). For example, in an *EGFR* L858R lung adenocarcinoma (Case #1), erlotinib was not responsive. WGS of the post-treatment tissue identified the ecDNA-driven hyper-amplification of EGFR reaching over 80 copies, which likely amplified the wild-type *EGFR* allele (**Figure 6B**), suggesting dual independent activating mechanisms of the MAPK pathway in the cancer (**Figure 6C**). It directly indicated that erlotinib inhibition which targeted only (*EGFR* L858R mutation) would not be sufficient.

From other seven cases, pathology review was insufficient to reveal the origin of tumors therefore selection of optimal treatments were challenging (**Category II-2**). Intriguingly, WGS was helpful for all the cases (7 out of 7; 100%). For example, Case #34 had a 6.4 cm jejunal mass, which was initially diagnosed as sarcomatoid carcinoma from unknown origin in pathologic evaluation (**Figure 6D**). Positron Emission Tomography (PET) scan discovered three metabolically active masses, including two lung nodules and one large adrenal mass. Application of WGS using the jejunal tumor revealed a heavy mutational load attributable to tobacco smoking (i.e., over 43,509 somatic mutations; 36.4% of base substitutions to SBS4; 38.7% of indels to ID3; **Figures 6E, 6F**). This data strongly suggests direct exposure of the cancer cell lineage to tobacco smoke ^53^, thus most likely primary origin is the lung, rather than adrenal gland or jejunum *per se*. Given the information, the cancer was diagnosed with non-small cell lung cancer with sarcomatoid differentiation that had metastasized into other organs, and then treated with the Checkmate-9LA regimen: nivolumab, ipilimumab, paclitaxel, and carboplatin, accordingly ^54^. The patient demonstrated remarkable and sustained responses (**Figure 6G**).

Finally, three cases benefited from WGS to evaluate their familial features (**Category II-3**). A colorectal cancer patient (Case #25) had first-degree relatives who had colorectal cancer (one of the parents) and a total colectomy (sibling) history. In WGS assay, a germline frameshift indel was found in the *APC* tumor suppressor gene, which was combined with loss-of-heterozygosity in cancer cells, which led to diagnose the patient as autosomal dominant familial adenomatous polyposis (**Figure 6H**) ^55^. Interestingly, WGS enabled to make an opposite conclusion in a similar circumstance. A breast cancer patient (Case #4) had bilateral breast masses which were metastasized to lung and pleura. Her sibling died from ovarian cancer at a young age (in her 50s), and her parents had stomach and cholangiocarcinoma history (**Figure 6I**). Clinically, the patient was strongly suspected as a familial breast cancer due to *BRCA1* or *BRCA2* pathogenic variants. However, no germline pathogenic variants were not found in WGS. In addition, the mutational patterns in cancer suggest that her breast cancer was a typical luminal type (**Figure 6J**) and HRD-associated mutational signatures were completely absent (HRD score = 0; **Figure 6K**), reassuring that the cancer was a sporadic case.

## Discussion

In this prospective hospital-based cohort study, we evaluated the clinical utility of WGS testing in precision oncology for solid tumors. A challenge in the clinical application of WGS is achieving standardized processing and the medically relevant interpretation of extensive sequencing data within the constrained timeframes necessary for clinical decision-making ^56^. This study utilized the commercially available CancerVision^TM^, a CLIA- and CAP-certified clinical assay. This assay incorporates a suite of automated, comprehensive bioinformatics pipelines that explore various dimensions of cancer genomes, including somatic point mutations, CNAs, SVs, germline pathogenic alterations, as well as analysis of mutational burden and signature. This approach allowed clinicians to implement a versatile, single-test solution in the clinical environment.

As demonstrated in this work, the assay spans a wide range of clinical questions, primarily informing on two main areas, i.e., seeking actionable genomic biomarkers (“actionability”) and providing clarity for clinical questions (“clarity”). From this comprehensive assay, patients can have a holistic view of their cancer. This includes identifying potential targeted treatment options, alignment with ongoing clinical trials, understanding of germline predispositions to cancer, and molecular insights that can be relevant for the potential of therapeutic resistance and pinpointing the cancer’s origin. Though not demonstrated in this study, WGS has broader applications, including evaluating precancerous lesions like polyps and monitoring residual diseases using tumor genomic data ^57,58^.

The definition of “clinical actionability” varies across studies, often confined to direct therapeutic implications. For instance, the REALM study (enriched with lung cancer cases) showed ∼40% clinical actionability from the TPS in on-label drug choices ^59^. Defining actionability in terms of therapeutic impact can be biased, as different cancer types have varying numbers of available treatments. For example, cancers like lung adenocarcinomas with more treatment options might show higher perceived utility in genomic tests. Despite the heterogeneity of our real-world cohort, our study echoes the previous observation, reporting an actionability rate of 37% for the same criteria. However, such a constrained view on actionability may not capture the full spectrum of individual tumor complexities. In our endeavor to provide a broader perspective on actionability, we evaluated the benefits of WGS for its comprehensive analysis of both somatic and germline alterations. In this cohort, we observed that WGS of tumor/normal samples provided clinically valuable insights in ∼70% of patients (67/95). It’s plausible that including additional aspects of utility, not covered in this study, could further enhance this actionability rate.

An indirect, but important benefit of the clinical application of WGS is the opportunities it creates for population-scale research from the clinical side. Globally, approximately 20 million new cancer cases are reported annually. For the last decade, academia-driven cancer genome studies explored at best 100,000 cancer whole-genomes worldwide, most of which lacking detailed medical history. These efforts have primarily identified frequent driver mutations from early stage, surgical tumor specimens, but the clinical impact of many of these mutations remain unclear. Cancer’s highly individualized nature, arising from diverse and heterogeneous genomic mutations, suggests that many less frequent driver mutations may go undetected ^60^. Particularly, mutations associated with later stages of cancer—such as recurrence, metastasis, and drug resistance—are likely more heterogeneous and rarer than those involved in initial tumorigenesis. To effectively capture and clinically annotate these mutations, systematic and comprehensive datasets are crucial. These datasets should encompass genome-wide profiling through WGS and complete clinical histories, covering a wide patient population (clinic-driven research). Genomic data from different hospitals need to be integrated without platform differences. These will be possible when WGS is widely adopted for the clinical routine procedures.

Until recently, integrating comprehensive tumor genome analysis into routine practice faced significant barriers, including the high cost of sequencing and the complexity of managing large data sets. As a result, concessions were often made. Genetic analyses were largely confined to exonic regions, with whole exome sequencing (**WES**) covering only about 1-2% of the entire genome. In some cases, the focus narrowed even further to targeted panel sequencing of the exonic regions of pertinent genes, representing a scant 0.3% of the genome. While economic studies indicate that using TPS testing can yield cost savings for both **CMS** (Centers for Medicare and Medicaid Services) and commercial payers when contrasted with standard limited gene testing ^61–63^. TPS approach can necessitate multiple tests, for example, for somatic, germline, and HRD analyses. This requirement can lead to additional costs being incurred. The WGS approach emerges as a more financially prudent option for in-depth genomic profiling. By deriving insights on germline, somatic, genomic instability, and mutation signatures from one single test, WGS not only ensures cost savings but also reduces diagnostic burden.

### Conclusion

The CancerVision WGS testing employed in this study, demonstrates its ease of use and clinical utility for solid tumor patients.

## Data Availability

All data produced in the present study are available upon reasonable request to the authors

## ACKNOWLEDGMENTS

We thank all the participating patients and their families and all the health care providers in Ajou University Hospital for their cooperation. Genome Insight supported this study in terms of the sequencing provision, and financial support. This research was supported by a grant of the Korea Health Technology R&D Project through the Korea Health Industry Development Institute (KHIDI), funded by the Ministry of Health & Welfare, Republic of Korea (grant number : HI23C1589 and HR22C1734 to Minsuk Kwon); the Bio & Medical Technology Development Program of the National Research Foundation (NRF) funded by the Korean government (MSIT) (NRF-2022M3A9G101451222 to Young Seok Ju); the new faculty research fund of Ajou University School of Medicine (Minsuk Kwon).

## AUTHOR CONTRIBUTIONS

R.K. (Ryul Kim), S.K. (Seokhwi Kim), B.B.L.O. (Brian Baek-Lok Oh), W.S.Y. (Woo Sik Yu), C.W.K. (Chang Woo Kim), Ho.H. (Hoon Hur), S.-Y.S. (Sang-Yong Son), M.J.Y. (Min Jae , ang), D.S.C. (Dae Sung Cho), T.H. (Taeyang Ha), S.H. (Subin Heo), J.Y.J. (Jeon Yeob Jang), J.S.Y. (Jae Sung Yun), K.-S.K. (Kyu-Sung Kwack), J.K.K. (Jai Keun Kim), J.H. (Jimi Huh), S.G.L. (Sun Gyo Lim), S.U.H. (Sang-Uk Han), H.W.L. (Hyun Woo Lee), J.E.P. (Ji Eun Park), C.-H.K. (Chul-Ho Kim), J.R. (Jin Roh), Y.W.K. (Young Wha Koh), D.L. (Dakeun Lee), J.-H.K. (Jang-Hee Kim), G.H.L. (Gil Ho Lee), C.K.N. (Choong-Kyun Noh), Y.J.J. (Yun Jung Jung), J.W.P. (Ji Won Park), S.S. (Seungsoo Sheen), M.S.A. (Mi Sun Ahn), Y.W.C. (Yong Won Choi), T.H.K. (Tae-Hwan Kim), S.Y.K. (Seok Yun Kang), J.-H.C. (Jin-Hyuk Choi), S.Y.B. (Soo Yeon Baek), K.M.L. (Kee Myung Lee), S.I.K. (Sun Il Kim), S.H.N. (Sung Hyun Noh), S.-H.K. (Se-Hyuk Kim), Hy.H. (Hyemin Hwang), E.J. (Eunjung Joo), Si.L. (Shinjung Lee), J.Y.S. (Jong-Yeon Shin), J.-Y.Y. (Ji-Young Yun), J.P. (Junggil Park), K.Y. (Kijong Yi), Y.K. (Youngoh Kwon), W.-C.L. (Won-Chul Lee), H.P. (Hansol Park), J.L. (Joonoh Lim), B.Y. (Boram Yi), J.K. (Jaemo Koo), J.-Y.K. (June-Young Koh), Sa.L. (Sangmoon Lee), Y.L. (Yuna Lee), B.-R.L. (Bo-Rahm Lee), E.C.-S. (Erin Connolly-Strong), Y.S.J. (Young Seok Ju), M.K. (Minsuk Kwon) Conceptualization: M.K., B.B.-L.O., Y.S.J. Project administration: M.K., B.B.-L.O., Hy.H., E.J.J., Si.L., Y.S.J. Supervised the patient enrollment and participated and handled study participants: W.S.Y., C.W.K., Ho.H., S.-Y.S., M.J.Y., D.S.C., T.H., S.H., J.Y.J., J.S.Y., K.-S.K., J.K.K., J.H., S.G.L., S.-U.H., H.W.L., J.E.P., C.-H.K., G.H.L., C.K.N., Y.J.J., J.W.P., S.S., M.S.A., T.H.K., S.Y.K., J.-H.C., S.Y.B., K.M.L., S.I.K., S.H.N., S.-H.K., M.K. Verify pathological specimen: S.K., J.R., Y.W.K., D.L., J.-H.K. Data curation: R.K., B.B.-L.O., Si.L. Methodology: J.Y.S., J.-Y.Y., Ju.P., K.Y., Y.O.K., W.-C.L. Acquisition, analysis, and/or interpretation: J.Y.S., H.P., J.L., B.Y., J.K., J.-Y.K., Sa.L., Y.L., B.B.-L.O. Supervision: B.B.-L.O., E.J.J., M.K, Y.S.J. Writing original draft: R.K., M.K, Y.S.J. Writing review and editing: R.K., S.K., M.K., B.B.-L.O., E.C.-S., Y.S.J.

